# Evaluation of Group Testing for SARS-CoV-2 RNA

**DOI:** 10.1101/2020.03.27.20043968

**Authors:** Nasa Sinnott-Armstrong, Daniel L. Klein, Brendan Hickey

## Abstract

During the current COVID-19 pandemic, testing kit and RNA extraction kit availability has become a major limiting factor in the ability to determine patient disease status and accurately quantify prevalence. Current testing strategies rely on individual tests of cases matching restrictive diagnostic criteria to detect SARS-CoV-2 RNA, limiting testing of asymptomatic and mild cases. Testing these individuals is one effective way to understand and reduce the spread of COVID-19.

Here, we develop a pooled testing strategy to identify these low-risk individuals. Drawing on the well-studied group testing literature, modeling suggests practical changes to testing protocols which can reduce test costs and stretch a limited test kit supply. When most tests are negative, pooling reduces the total number of tests up to four-fold at 2% prevalence and eight-fold at 0.5% prevalence. At current SARS-CoV-2 prevalence, randomized group testing optimized per country could double the number of tested individuals from 1.85M to 3.7M using only 671k more tests.

This strategy is well-suited to supplement testing for asymptomatic and mild cases who would otherwise go untested, and enable them to adopt behavioral changes to slow the spread of COVID-19.

## Introduction

Group testing was first described in [5] as a technique for screening United States Army recruits for syphilis. In a common extension, samples are organized into a matrix, then each row and column of the matrix are tested [13, 6]. Under either approach, it is well understood that given a sensitive test and low disease prevalence, testing capacity increases dramatically.

We propose adapting this technique for diagnosis of SARS-CoV-2, where COVID-19 prevalence is low and testing is limited. This is in part motivated by work suggesting that population-scale testing for SARS-CoV-2 might effectively reduce asymptomatic transmission [1]. During the past week, as we finalized this manuscript, its application to SARS-CoV-2 testing has been widely suggested and demonstrated [20, 19, 8].

Since group testing is most efficient when prevalence is low, we propose using it to test individuals who do not meet current test criteria. This obviates many concerns around group testing efficacy on the individual level, as critical care resources are not essential for asymptomatic cases. Individuals with a high case probability can be excluded from groups and tested individually using either existing criteria for screening or newly developed survey instruments [16, 17].

While CDC guidelines [4] caution against using negative test results “as the sole basis for treatment or other patient management decisions,” testing criteria currently select for symptomatic individuals. Among individuals presently excluded by testing guidelines, we argue that grouped testing is an effective strategy for finding asymptomatic cases that would otherwise be missed.

Overall, we propose a very simple pooling strategy (Figure 1) and thoroughly evaluate the relevance of this test to the COVID-19 pandemic. First, on a 96-well plate, pool each row (8) and each column (12), and perform a total of 20 tests on the pooled samples across the 96 individuals being tested (plus appropriate positive and negative controls). Then, all samples for which both the row and column were positive are re-run in a validation test to determine which individuals are positive. We show below that this strategy works to improve upon existing testing and evaluate the benefits across both simulated and real testing data.

**Figure 1:**
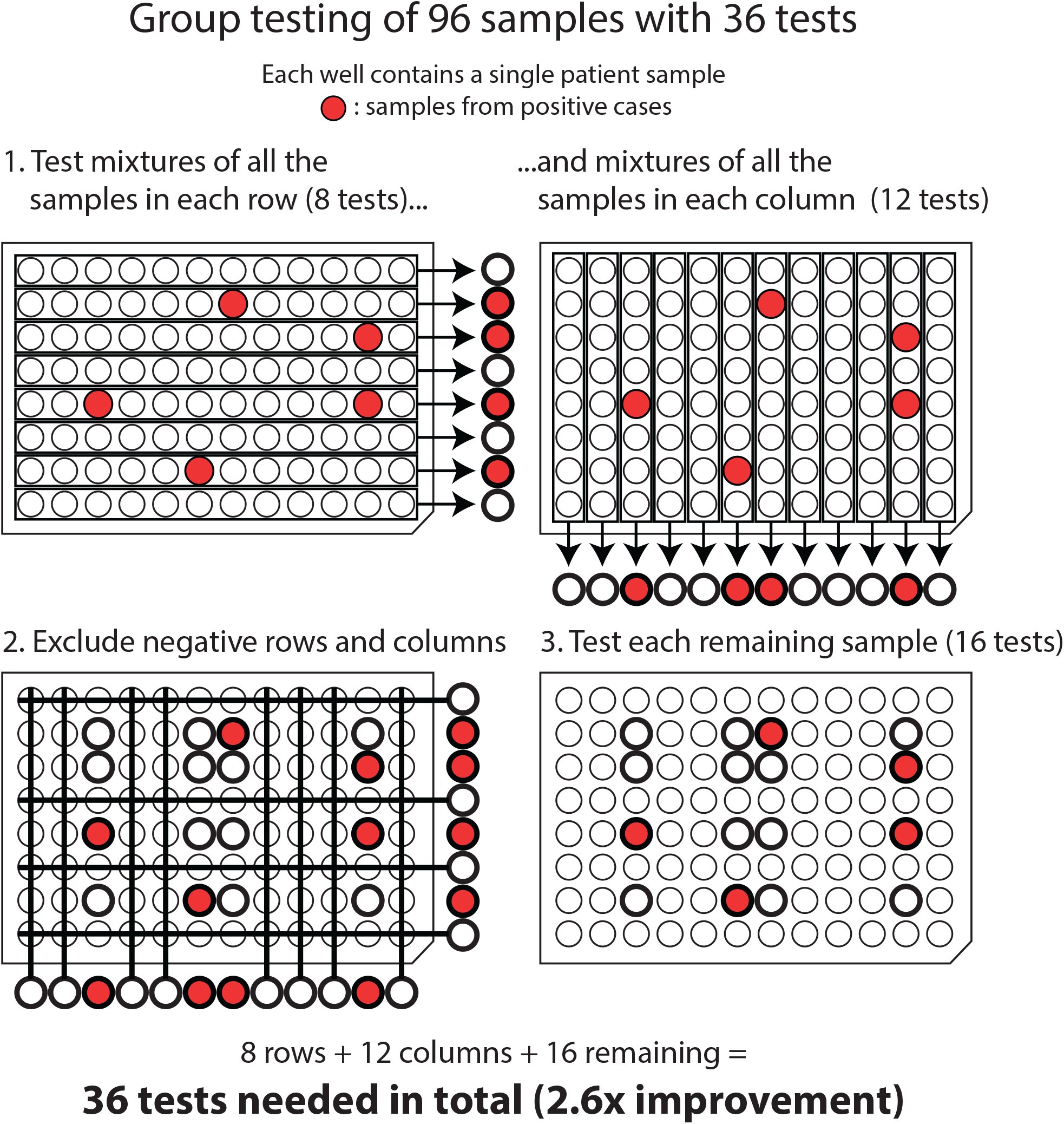
Overview of the testing procedure. On a 96-well plate, pools of samples from each row, and separate pools of samples from each column, are tested in a first round. Positive rows and columns are present if any individuals therein are positive, so negative rows and columns can be excluded. The individuals from positive rows and columns are then tested individually to assess their COVID-19 status. Individuals for which both their row and column tested negative are not screened further. An interactive tool is available at https://technopolymath.shinyapps.io/pooled_covid_testing/

## Results

First, we evaluated whether group testing might improve upon existing individual-test methodologies. Testing three pooling strategies — four-well pooling (pooling four adjacent wells in a single dimension), 96-well (8×12) plate column and row pooling, and 384-well (16×24) plate column and row pooling — we found that all improve significantly upon naive testing, and that each performed better for a particular prevalence, as expected (Figure 2). All three required fewer tests than the naive method at prevalence less than 10%. We have implemented these simulations in an R package that is available at https://gitlab.com/technopolymath/pooled-diagnostics, and an interactive web visualization is available at https://technopolymath.shinyapps.io/pooled_covid_testing/.

**Figure 2:**
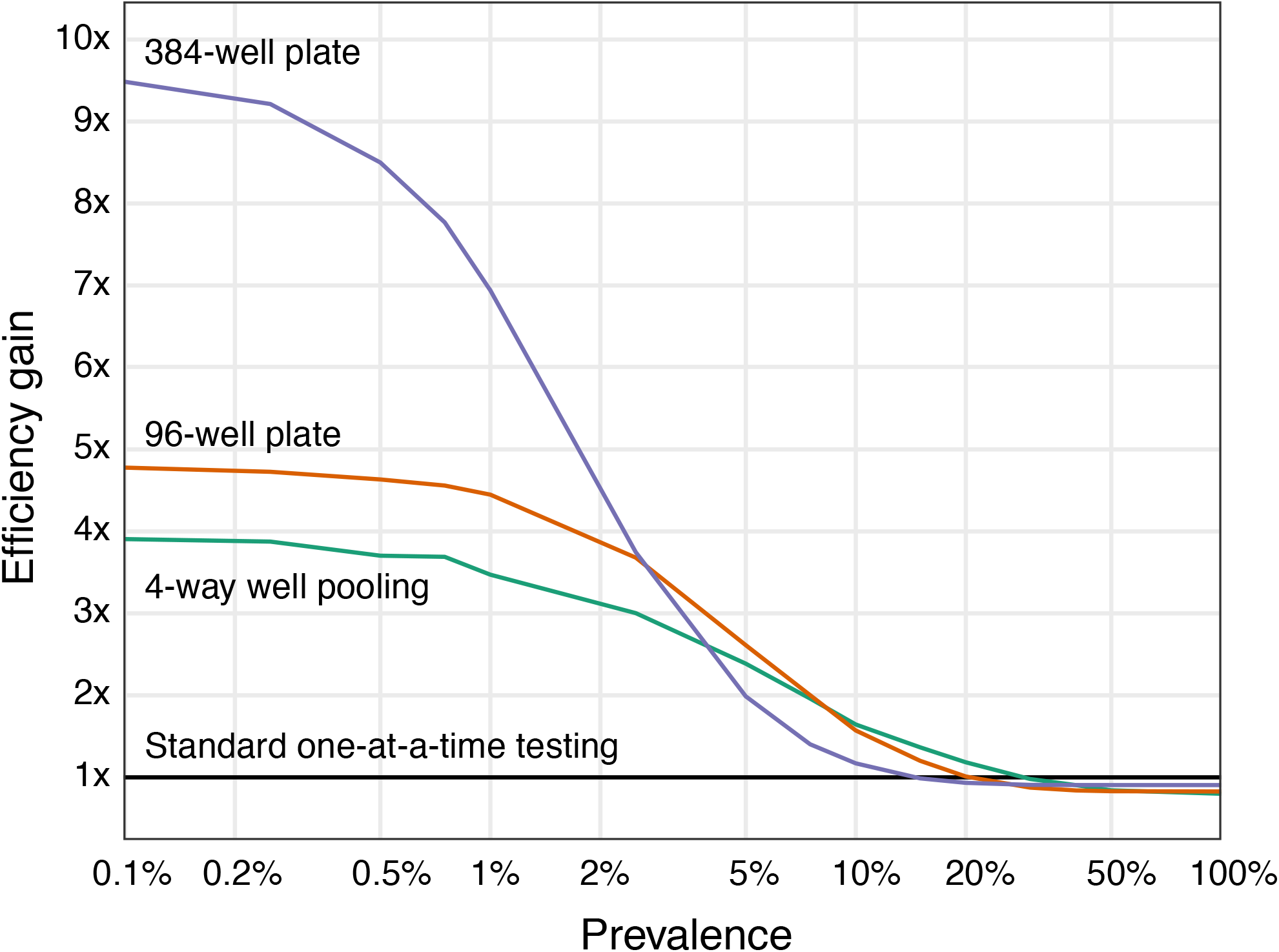
Evaluation of mean improvement of the pooling strategies relative to naive testing under different predicted prevalence. At 2% prevalence and below, row and column pooling on 384-well plates performs best; between 2% and 10%, row and column pooling on 96-well plates performs best; and at greater than 10% prevalence, four-way pooling of wells performs best.

We additionally wanted to estimate the total saved tests relative to those conducted so far, under the hypothetical that pooling had been used throughout the pandemic. Using testing and positive test estimates from Our World in Data [15], we chose an optimal testing strategy per country from between 4-well pooling; 96-well plates; and 384-well plates. We chose the strategy per country which minimized the tests needed at the observed prevalence. This resulted in an expected total of 670,849 tests globally to re-test everyone who has previously been screened for SARS-CoV-2 (1.85M individuals), corresponding to a three-fold improvement in cases per test. Ignoring country-specific prevalence data, 96-well plates and between 5-way and 8-way well pooling resulted in an expected ≈ 780, 000 tests (Table S1). When considering country-specific prevalence, however, the single strategy which performed best on average across countries was the 96-well plate (at just under a third as many tests as standard approaches; Figure S3). These findings suggests that the 96-well plate row and column pooling approach is a straightforward compromise if no information on population prevalence is available.

This approach is even better suited to mild and asymptomatic individuals, as population prevalence is most likely substantially lower than observed in the cases tested by most countries, an intuition borne out in recent modeling efforts [14]. We assume that the prevalence estimates from existing testing are biased due to primarily testing symptomatic individuals, and therefore evaluated our approach at a range of prevalence biases (Figure S2). At up to ten-fold lower prevalence than current tests, our approach still discovers more cases per test than existing diagnosis protocols.

Together, these results suggest that testing mildly symptomatic, or asymptomatic, individuals using a group testing approach will discover the same number of cases using the same number or fewer tests, albeit in a population with much lower prevalence. This enables large numbers of lower-risk individuals to be tested, either from mild symptomatic individuals or contact tracing, which in turn empowers behavioral change.

## Discussion

We propose a pooling extension to existing SARS-CoV-2 testing protocols to boost efficiency and better utilize scarce resources. These are not meant to be a replacement for wide-spread testing, but instead might act as a stop-gap measure to reduce the number of RNA extractions and testing steps performed while still identifying asymptomatic or mild cases. Even so, there are numerous reasons why the use of such strategies is not popular.

1. Grouped testing, particularly with an iterative test as here, uses more consumables, such as pipette tips, than traditional methods per test. This is at the benefit of more individuals being tested.
2. More complex sample assignment strategies (e.g. probability driven) can reduce the total number of tests but these are difficult to implement in practice. As such, we frame our proposal in terms of the standard 96-well plates and show that a single plate’s worth of pooling already is very effective at expected population prevalence, supporting the use of this strategy (Figure S4).
3. Similarly, the random assignment of individuals to wells, which is much easier than clinical-risk-driven ordering, has nearly the same improvement (Figure S1). In addition, the total improvement in tests between group testing everyone and only group testing low-risk individuals is nearly indistinguishable at testing prevalence below 10%, suggesting that either is an equally useful option to pursue.
4. Mixing samples from multiple individuals in a single tube risks contamination. We reiterate that this method should not be used as the sole diagnostic criterion for critical ill patients, but rather can be considered to develop epidemiological parameters and/or perform initial screening in low-risk individuals then follow up with confirmatory tests.
5. Interpretation and reporting requirements assume individual testing. In particular, CDC guidelines [4] requires a human RNA positive result (RP+) for an interpretation of “2019-nCoV not detected”. In the pooled strategy, validation tests will be run on a number of negative cases, which may be sufficient to demonstrate a high RP+ rate on this population. Pooled negatives might be considered as “Invalid Results”, and thus should not be reported, until such time as RP+ is confirmed individually.
6. Picking individual samples to re-test is prone to pipetting errors. We developed a strategy with this in mind, as it only requires re-testing samples from any individual row or column which tests positive as a pool. In this way, the workflow disturbance is minimized. See “Minimizing pipetting errors” below.
7. If RNA extraction is the limiting step, this approach requires samples to stay stable for the first qPCR run, in order to allow RNA extraction from the positive samples. We note there is some evidence that RNA extraction is not necessary for qPCR of SARS-CoV-2 RNA [3], though more testing is needed.
8. This method does not remove any clinical or logistical barriers to obtaining large numbers of individual swabs for testing. As such, it relies on the presence of other infrastructure to obtain prevalence estimates. Alternative strategies for prevalence-only modeling, such as wastewater-based epidemiology [7], might be more suitable in settings where individual swabs are not available.
9. At non-negligible prevalence, a substantial number of tests are still needed to cover large populations. The four-fold improvement we observe at prevalence 2% still results in large numbers of total tests, which might remain above capacity.
10. Requiring two stages of qPCR might delay return of results. However, testing resource limitations are also causing delays in the current pandemic, and confirmatory tests are often employed for diagnostics. Finally, for the targeted subset of people who have COVID-19 but do not have severe symptoms, delays may have limited clinical significance or consequences for public health interventions.

We encourage further work into better approximate methods for obtaining prevalence estimates through more sophisticated pooling strategies, including using the virus concentration from qPCR (in either equally or unequally pooled samples). In addition, we see great potential of pooling for repeat testing of the same population of individuals on sequential days or weeks. Hospitals, government offices, corporations, and educational institutions, across which many individuals cannot work from home [2], require self-isolation following positive results. This decreases the case counts below the population average on subsequent days and increases the efficiency of group testing strategies. Among individuals with low risk of serious complications, this methodology could use non-qPCR based assays. For instance, RT-LAMP assays [9, 18, 21, 10] might be particularly well-suited to rapid, first-line screening methodologies at scale. This strategy might also be applied to seropositivity screening once reliable tests are available for COVID-19, though rigorous testing of the pooling of serum is necessary to evaluate this more thoroughly.

Further extensions of this approach might be justifiable given the current extreme limitations on testing resources. For instance, testing efficiency can be increased at the expense of specificity by pooling people in close contact (ex. family groups, work units) into a single sample prior to group allocation or RNA extraction. This reduces test volume by a constant factor under the assumption that individuals in close contact have positively correlated infection status and that cluster identification is more vital to suppression than determining a particular individual’s infection status. In addition, pooling samples from the same individual (sputum, nasopharyngeal swab, etc) will obtain similar benefits with essentially no loss of information.

Overall, we present a straightforward and directly applicable approach to extend testing for SARS-CoV-2 to lower-risk individuals using pooling. While there are numerous technical hurdles left to overcome, we anticipate that strategies such as those proposed here will be valuable in obtaining more accurate estimates of population prevalence and disease spread in the coming months.

### Minimizing pipetting errors

Pipetting errors are clearly a large concern in the implementation of this approach, as single missteps are both difficult to detect and result in both false positives and false negatives. We consider multiple possible mitigation strategies:

1. Multi-channel pipettes enable simultaneous multi-sample manipulation by hand. Given the gridded nature of our testing system, multi-channel pipettes are likely effective. Multi-channel pipetting into a deepwell plate for pooled sample storage is likely the simplest option. For example, into a 2.5 mL deepwell plate, pipette 150 *µ*L of each pre-extraction row and column sample to yield a total of 1.8 mL of pool for rows and 1.2mL of pool for columns. Then mix thoroughly and extract from standard volumes. We note that this mixing increases risk of operator exposure, so strict biosafety precautions must be adhered to.
2. Many SARS-CoV-2 testing laboratories are using pipetting robots to rapidly and accurately scale RNA extractions and assays to large numbers of individuals. While protocols do not exist to our knowledge for group testing on these platforms, they are ideally suited to tasks such as this. We propose such a strategy should be validated using traditional pipetting before using valuable robotic platform resources.
3. Finally, in the absence of both robotic and multi-channel automation, pipette guiding tools such as iPipet [22] can be used for suggesting pipetting steps in a fixed order. This reduces the possibility of pipetting errors without requiring a multi-channel pipette, but does require tablet access.

Independent of the modest efficiency gains of clinical-risk-driven ordering (as in Figure S1), ordering of individual samples so that pipetting order is from low-to high-risk samples minimizes the impact of unintended transfers. In the example strategy in Figure 1, this would be achieved by placing high-risk samples in the bottom-right.

## Data Availability

This is a work of secondary analysis and all data are available from other studies or websites. An interactive simulation framework derived from the work presented in this study is available at https://technopolymath.shinyapps.io/pooled_covid_testing/.

https://ourworldindata.org/covid-testing

## Acknowledgments

We would like to thank Xinchen Wang, Hanna Ollila, Diego Calderon, Courtney Smith, Josh Tycko, Veronica Casas, Michael Wainberg, Jonathan Pritchard, Ruth Brillman and Christopher Davis for giving comments on this work. In particular, Courtney Smith suggested the use of pooling to evaluate individuals post infection, Michael Wainberg provided advice on figure design, and Xinchen Wang and Diego Calderon altered us to recent references. N.S.-A. is supported by a Stanford Graduate Fellowship. We thank Our World in Data for providing and updating the global testing effort data. We thank numerous government agencies for providing up-to-date information on COVID-19.

## Supplement

We provide an overview of the simulation methods employed. We evaluated models using a sampling approach, drawing samples from either a Bernoulli distribution with a probability equal to the assumed prevalence, or a beta-Bernoulli distribution with *α* parameter 5 and *β* parameter 5*/prevalence* to account for inter-individual variation in risk. All draws were independent. These were averaged over 1000 replicates to smooth over any sampling noise; sample sizes are adjustable in our web interface.

Next, the draws are used to assign true case status for each simulation run. The number of random values thus corresponds directly to the count of cases tested by a given method in a single batch. Of note, the draws from the beta are used as the case probability directly for the row major ordered case prevalence tests (Figure S1), which are only informative under the beta-Bernoulli simulations.

Finally, the results with our two-stage approach (pooled screening followed by individual-test followup of potential cases, and optional confirmation of positive status) are compared with corresponding naive screening (individual-test screening followed by optional and default individual-test confirmation of cases). Evaluating different levels of test stringency for both the grouped and naive strategies is part of the web interface.

We assume for all methods that samples are distributed at random and independently. However, sample collection for this approach might be conducted through contact tracing and door-to-door sampling, where we expect co-habitation. In these cases, the assumption of lack of correlation of test status between individuals does not hold. We anticipate relevant extensions to this work might address this concern.

While the random assignment might at first seem inefficient, we evaluated a number of additional strategies, including ones which use the case probability information, and found that they did not perform meaningfully better than random assignment (Figure S1). As such, given the ease of the random assignment process, we recommend that as a default. Comparison to case ordered alternatives is available on the web interface.

Strategies that pool across multiple plates offer further test efficiency gains, depending on the prevalence. Because this dependence is sensitive to prevalence and because using a single 96-well plate is near-optimal at an expected 5% prevalence (Figure S4), we recommend this practically simpler strategy.

Sample stratification provides another opportunity for realizing efficiency gains. By separating samples from symptomatic individuals from others, we can artificially decrease prevalence in the individuals tested using pooling, which decreases the tests per individual. This makes it easier to find asymptomatic carriers by expanding test coverage, though practical trade-offs between case abundance and overall testing capacity are worthy of consideration.

**Figure S1:**
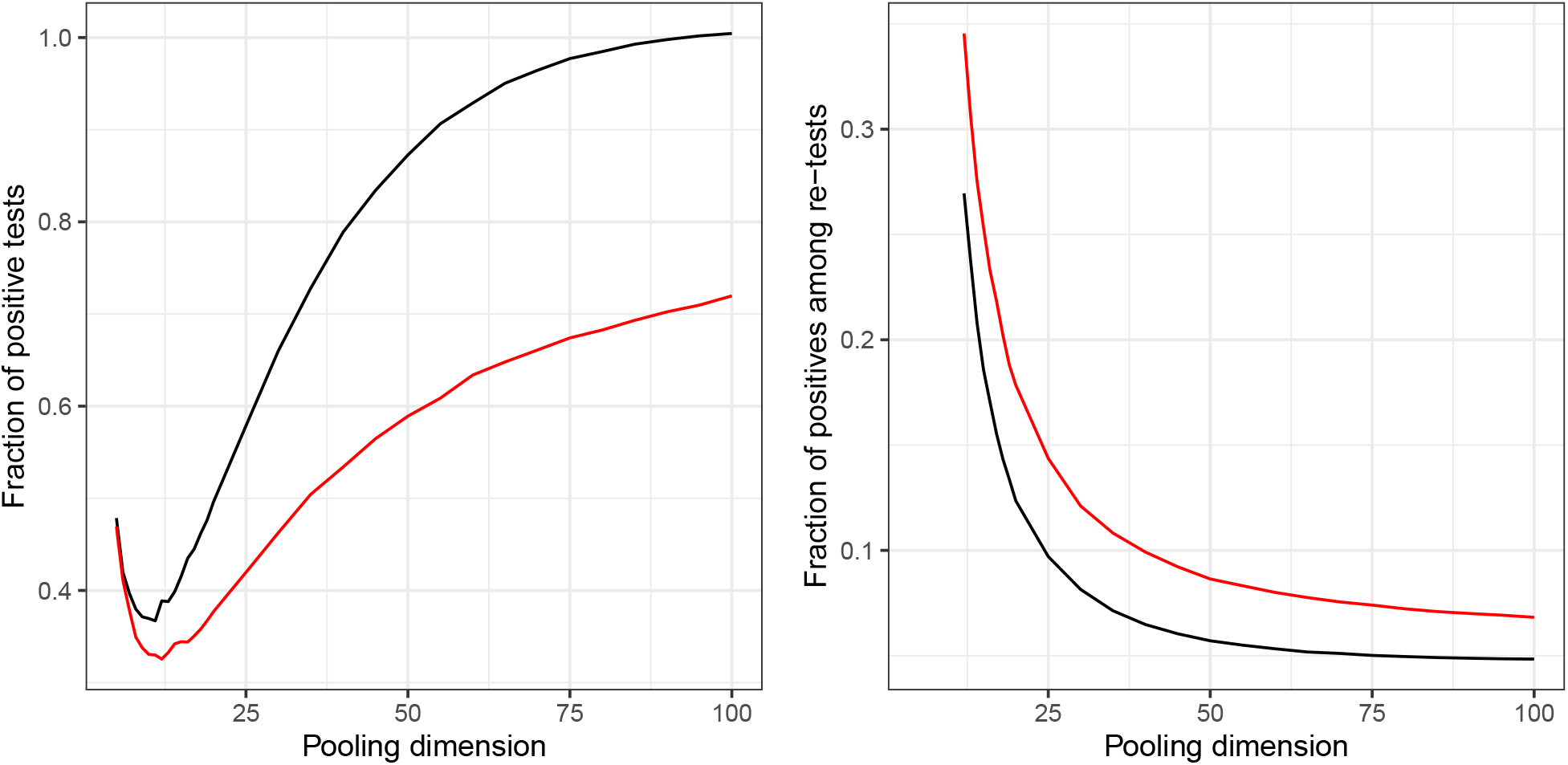
Comparison of random (black) versus case-probability-ordered (red) modes of pooling. Simulations done with known case probability drawn from *Beta*(0.5, 10) (5% prevalence). Pooling dimension *k* is the size of the *k* × *k* plate of individuals to be tested (rows and columns). In both the total number of tests relative to naïve (left) and the fraction of positives in the resulting secondary testing scheme (right), the ordered method results in slight improvements around the optimum pooling dimension. We suggest these are not sufficient to warrant the additional complexity this method entails.

**Figure S2:**
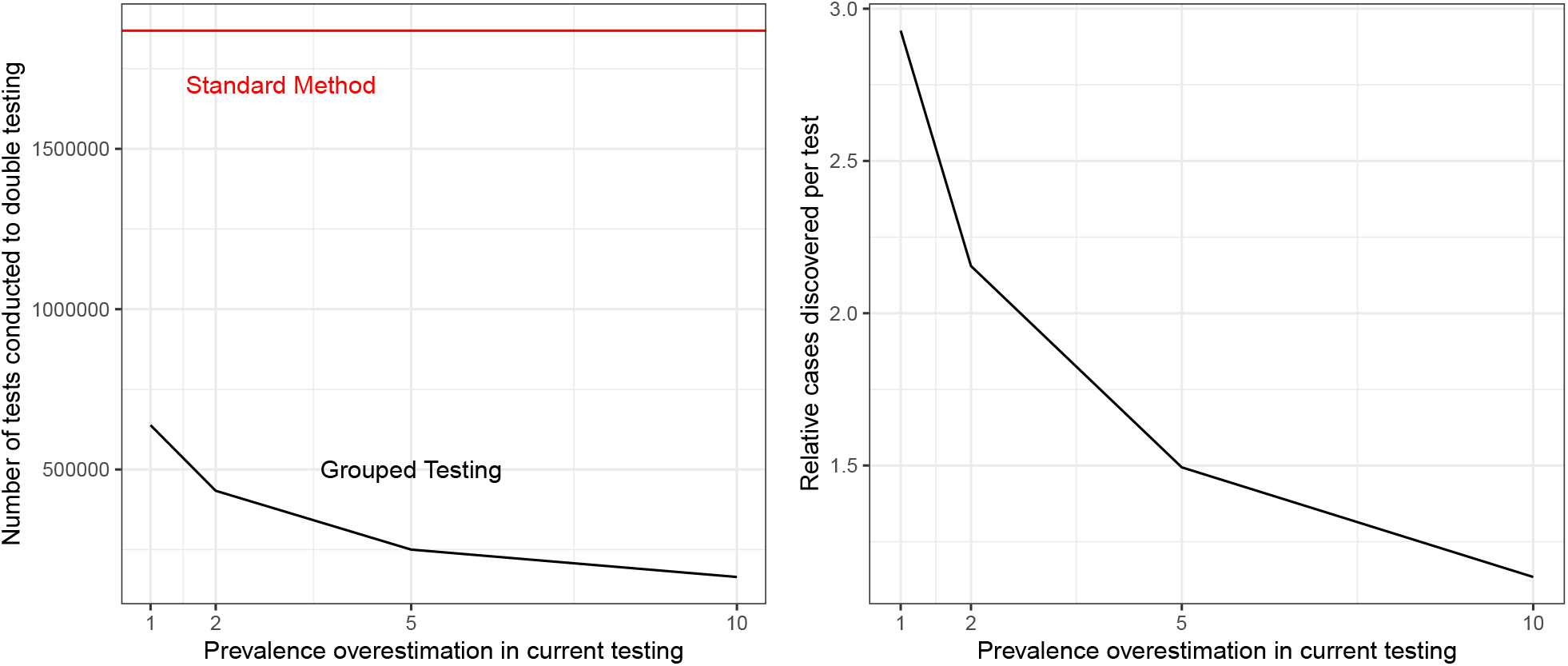
Expected improvements relative to current testing. Across global tests performed so far, we choose the best grouped testing strategy per country. In order to double the current tests performed (left), grouped testing requires progressively fewer total tests at lower prevalence. Prevalence scaled by observed prevalence within each country (so 2-fold overestimation in a country with 3% observed prevalence = 1.5% tested prevalence). Testing these lower-prevalence individuals using group testing finds more cases per test than existing strategies did, even at lower overall prevalence than existing screening (right).

**Figure S3:**
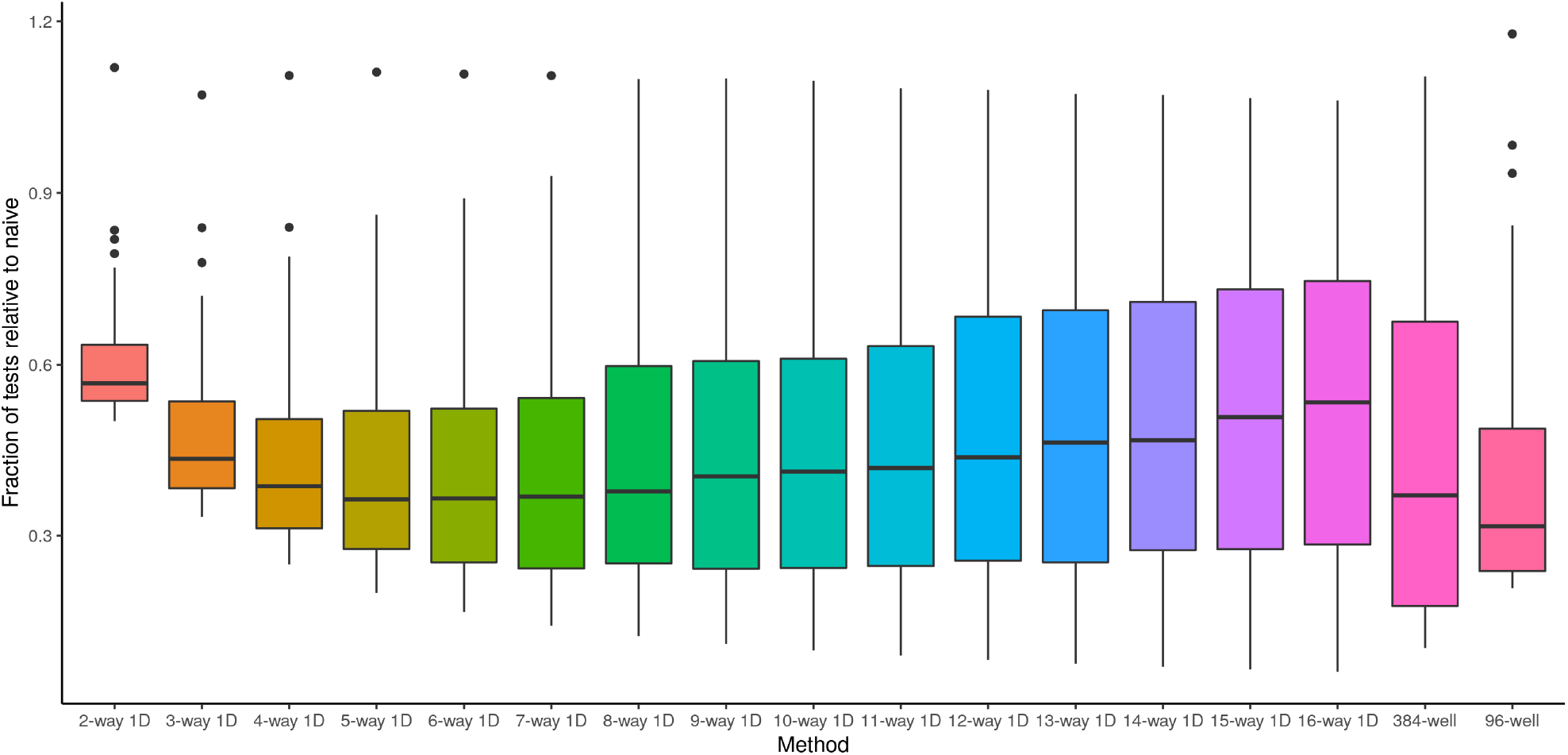
Comparison of pooling strategies across countries used in the optimization simulation. Across all 60 sampled countries [15], the 96-well plate row and column approach had the largest median improvement (just greater than three-fold). Alternative strategies that work well across a large number of countries are 384-well plates and 5-way to 8-way well pooling.

**Figure S4:**
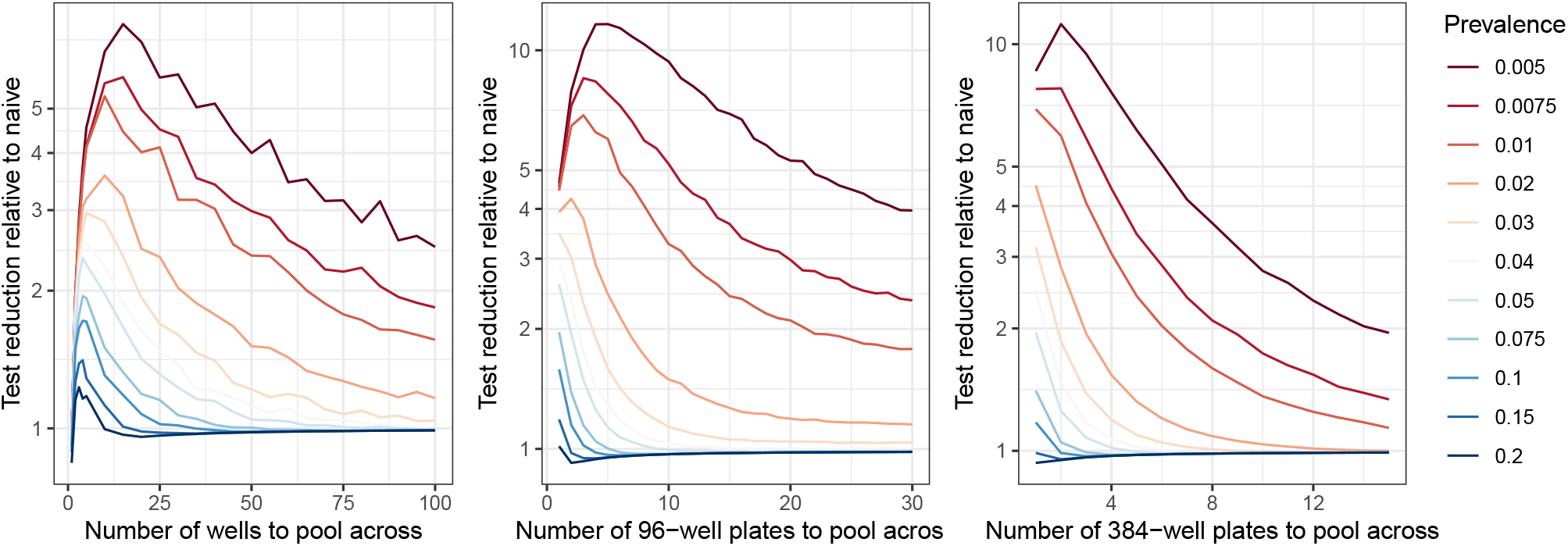
Comparison of prevalence scaling of efficiency for simple well pooling (left), 96-well plates (middle), and 384-well plates. Simple well pooling is more efficient at prevalence ≥ 10%, while 384-well plates are more efficient for prevalence ≤ 1%. Pooling of 3-5 wells and single 384-well plates is recommended at prevalence outside the range that is well captured by a single 96-well plate.

**Table S1:**
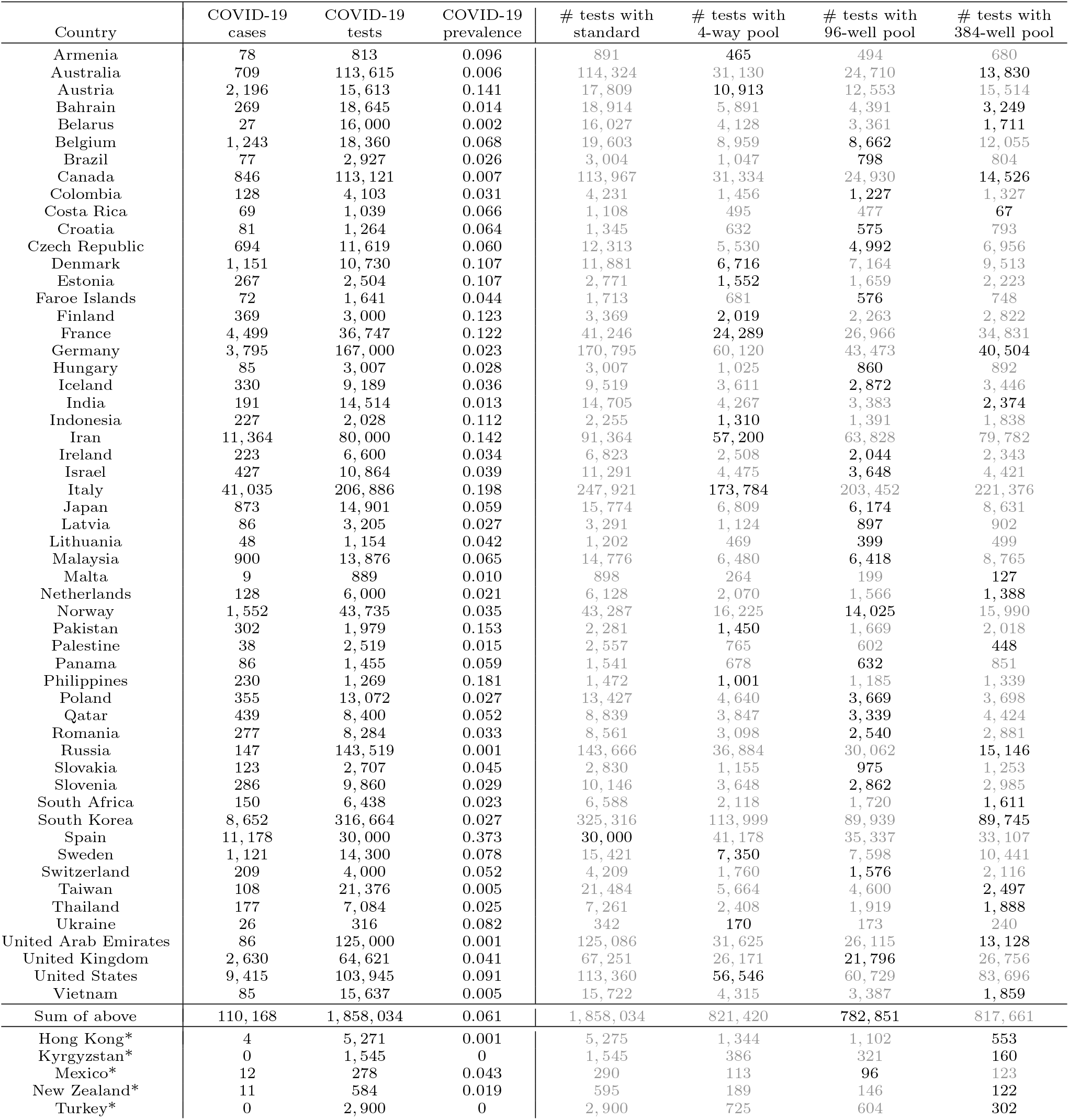
Comparison of pooling strategies across countries and choice of the 96-well plate as a recommended strategy. Across 55 countries with available matched case and testing data [15], the estimated number of tests required to duplicate the existing testing effort (at country-observed prevalence among tested individuals) exclusively using the given method is provided. When examining how many countries are optimized by each strategy, however, 40% (24 of 55) of countries save the most tests using the 96-well plate strategy. Choosing the best strategy per country results in a total of 670,849 tests, the vast majority of which are saved by the 384-well plate and 96-well plate strategies. Data from different countries are across a range of dates (3/5-3/20) as number of tests are not systematically reported. * Case counts were not recorded by [15] on the same day as test counts for Turkey, Kyrgyzstan [12], Hong Kong, Mexico [11], and New Zealand, are obtained from source and [15], and are excluded from global estimates. We infer zero cases in Turkey from the date reporting tests (3/10/2020) and first case date (3/12/2020) and exclude two probable cases in New Zealand.

## References

[1] Andrea Crisanti and Antonio Cassone. In one Italian town, we showed mass testing could eradicate the coronavirus. https://www.theguardian.com/commentisfree/2020/mar/20/eradicated-coronavirus-mass-testing-covid-19-italy-vo [Accessed: 2020-03-24]. March 20, 2020.

[2] Marissa G Baker. “Characterizing occupations that cannot work from home: a means to identify susceptible worker groups during the COVID-19 pandemic”. Mar. 2020.

[3] Emily A Bruce et al. “RT-qPCR Detection of SARS-CoV-2 RNA from patient nasopharyngeal swab using QIAGEN RNEeasy kits or directly via omission of an RNA extraction step”. In: bioRxiv (2020).

[4] Centers for Disease Control and Prevention, Division of Viral Diseases. CDC 2019-Novel Coronavirus (2019-nCoV) Real-Time RT-PCR Diagnostic Panel. https://www.fda.gov/media/134922/download [Accessed: 2020-03-23]. March 15, 2020.

[5] Robert Dorfman. “The detection of defective members of large populations”. In: The Annals of Mathematical Statistics 14.4 (1943), pp. 436–440.

[6] Yaniv Erlich et al. “DNA Sudoku - Harnessing high throughput sequencing for multiplexed specimen analysis”. In: Genome Research 19.7 (2009), pp. 1243–1253.

[7] Iria González-Mariño et al. “Spatio-temporal assessment of illicit drug use at large scale: evidence from 7 years of international wastewater monitoring”. In: Addiction 115.1 (Jan. 2020), pp. 109–120.

[8] Paul Hammel. .@GovRicketts says state is ‘pooling’ tests now, so combining five tests into one test tube. If it comes back ‘negative,’ then you know all were negative; if one was ‘positive,’ then those five will have to be retested. So that expands capacity to 400+ tests a day by public labs. https://twitter.com/PaulHammelOWH/status/1242539326568902661 [Accessed: 2020-03-24]. March 24, 2020.

[9] Minghua Jiang et al. “Development and validation of a rapid single-step reverse transcriptase loop-mediated isothermal amplification (RT-LAMP) system potentially to be used for reliable and high-throughput screening of COVID-19”. In: medRxiv (2020).

[10] Laura E Lamb et al. “Rapid Detection of Novel Coronavirus (COVID-19) by Reverse Transcription-Loop-Mediated Isothermal Amplification”. In: medRxiv (2020).

[11] Andrea Navarro. “Mexico’s Low Coronavirus Count Spurs Doubts On Testing Rate”. In: Bloomberg News (Mar. 2020).

[12] Maria Orlova. Government of Kyrgyzstan announces number of coronavirus test systems. https://24.kg/english/146493_Government_of_Kyrgyzstan_announces_number_of_coronavirus_test_systems_/. Accessed: 2020-3-26. Mar. 2020.

[13] RM Phatarfod and Aidan Sudbury. “The use of a square array scheme in blood testing”. In: Statistics in Medicine 13.22 (1994), pp. 2337–2343.

[14] Lionel Roques et al. “Mechanistic-statistical SIR modelling for early estimation of the actual number of cases and mortality rate from COVID-19”. Mar. 2020.

[15] Max Roser, Hannah Ritchie, and Esteban Ortiz-Ospina. “Coronavirus Disease (COVID-19)–Research and Statistics”. In: Our World in Data (2020).

[16] Hagai Rossman et al. “A framework for identifying regional outbreak and spread of COVID-19 from one-minute population-wide surveys”. In: medRxiv (2020).

[17] Arni S Srinivasa and Jose A Vazquez. “Identification of COVID-19 Can be Quicker through Artificial Intelligence framework using a Mobile Phone-Based Survey in the Populations when Cities/Towns Are Under Quarantine”. In: Infect. Control Hosp. Epidemiol. (2020), pp. 1–18.

[18] Mohamed El-Tholoth, Haim H Bau, and Jinzhao Song. “A Single and Two-Stage, Closed-Tube, Molecular Test for the 2019 Novel Coronavirus (COVID-19) at Home, Clinic, and Points of Entry”. In: (2020).

[19] Idan Yelin. Pool testing for COVID-19 works! Can detect a single positive sample within a pool of 64 samples. Collaboration of: @RambamHCC @TechnionLive @BiologyTechnion @RoyKishony @yuval geffen. https://twitter.com/idanyelin/status/1239930847748227079 [Accessed: 2020-03-23]. March 17, 2020.

[20] Idan Yelin et al. “Evaluation of COVID-19 RT-qPCR test in multi-sample pools”. In: medRxiv (2020). doi: 10.1101/2020.03.26.20039438. eprint: https://www.medrxiv.org/content/early/2020/03/27/2020.03.26.20039438.full.pdf. url: https://www.medrxiv.org/content/early/2020/03/27/2020.03.26.20039438.

[21] Lin Yu et al. “Rapid colorimetric detection of COVID-19 coronavirus using a reverse transcriptional loop-mediated isothermal amplification (RT-LAMP) diagnostic platform: iLACO”. In: medRxiv (2020).

[22] Dina Zielinski et al. “iPipet: sample handling using a tablet”. In: Nature methods 11.8 (2014), p. 784.

